# Risk of opioid-related mortality associated with buprenorphine versus methadone: A systematic review of observational studies

**DOI:** 10.1101/2023.08.13.23294034

**Authors:** Jihoon Lim, Imen Farhat, Antonios Douros, Soukaina Ouizzane, Dimitra Panagiotoglou

**Affiliations:** Department of Epidemiology, Biostatistics, and Occupational Health, McGill University, Montreal, QC, Canada; Department of Medicine, McGill University, Montreal, QC, Canada; Centre for Clinical Epidemiology, Lady Davis Institute, Jewish General Hospital, Montreal, QC, Canada; Institute of Clinical Pharmacology and Toxicology, Charité - Universitätsmedizin Berlin, Berlin, Germany

**Keywords:** Opioid agonist treatment, Mortality, Buprenorphine, Methadone, Comparative effectiveness research

## Abstract

**Introduction:** Buprenorphine and methadone are effective treatments of opioid use disorder (OUD) and can reduce drug-related mortality. While observational studies have compared head-to-head buprenorphine and methadone, this evidence has not been previously synthesized. Our study aims to systematically review the available evidence on the comparative effectiveness of buprenorphine and methadone in people with OUD, thereby rigorously assessing the methodological quality of individual studies.

**Methods:** We searched Medline, Embase, PsycINFO, and Web of Science for all relevant articles published between 1978 and April 8, 2023. Observational studies directly comparing the risk of drug-related mortality between buprenorphine and methadone among people with OUD were eligible. We assessed the overall risk of bias using the Risk Of Bias In Non-randomized Studies of Interventions (ROBINS-I) tool.

**Results:** Our systematic review included seven studies. There was mixed evidence of comparative mortality risk, with heterogeneity across study region, time, and treatment status (on treatment vs. discontinued). Three studies reported no difference, and four reported findings in favour of buprenorphine. Based on ROBINS-I, three studies had a moderate risk of bias, two had a severe risk, and two had a critical risk. Major sources of biases were residual confounding and selection bias along with presence of prevalent user bias, informative censoring, and left truncation.

**Conclusions:** Due to methodological limitations of the observational studies, generalizability of their findings remains unknown. Therefore, to provide a more accurate comparative safety profile for these two medications, further observational studies with methodological rigour are warranted.

## 1. Introduction

Opioid use disorder (OUD) is a chronic disorder characterized by problematic patterns of opioid use, potentially leading to serious adverse events including death. Among patients with OUD, drug-related deaths are the leading cause of mortality [1]. In 2021, the United States (US) and Canada reported 107573 and 8006 opioid overdose deaths, respectively [2, 3]. Opioid agonist treatments (OAT) such as buprenorphine or methadone are an effective pharmacotherapeutic intervention to treat OUD [4–9]. OATs demonstrate better treatment retention, reduce illicit opioid use, and lower overdose and all-cause mortality risks compared with detoxification and psychological treatment for OUD [10, 11]. Thus, they are recommended as first-line treatment for OUD [12–15].

Although overdose on methadone or buprenorphine is rare [16, 17], misuse and diversion, including the use of these drugs as substitutes for other drugs and self-medication, remain possible [18–20]. Studies have documented occurrences of fatal overdose or poisoning, but most cases involving these OATs indicate polysubstance use with other illicit drugs [21–24].

While both buprenorphine and methadone carry risks of adverse effects, evidence on their comparative safety remains unclear. For example, one Canadian study showed a higher frequency of detection of methadone than buprenorphine in fatal overdose cases [25]. On the other hand, a US-based study revealed a lower rate of opioid-related mortality associated with methadone than with buprenorphine among individuals who had experienced a non-fatal overdose [7]. Other studies have shown that treatment retention and completion rates appeared to be higher for methadone [26–28], but a recent review of 19 cohort studies showed a lower pooled overdose mortality rate attributable to buprenorphine [6]. Relatedly, suppression of illicit opioid use did not differ between buprenorphine and methadone [29, 30].

In light of the ongoing overdose epidemic, public health stakeholders have sought to expand access and improve retention to OAT [4, 14, 31]. Several observational studies have attempted to elucidate the comparative safety and risks of fatality associated with OATs, but earlier systematic reviews in this literature have focused on indirect comparison of mortality risks associated with buprenorphine and methadone [6, 32]. These indirect comparisons involved calculation of mortality rates during OAT (i.e., “on” treatment) and when not on treatment (i.e., “off” treatment) for buprenorphine and methadone separately, followed by comparison of these rates during exposure to each medication against those during the ‘off’ period. Such comparisons may not fully account for the heterogeneity in study population, study definition, and study design. In addition, these reviews have inadequately assessed the methodological strengths and limitations of individual observational studies in their evidence synthesis.

With this background, our objective is to systematically review the available evidence on head-to-head comparisons estimating the risk of drug-related poisoning mortality associated with buprenorphine or methadone in people undergoing treatment for OUD, and to rigorously assess the methodological quality of individual studies. To our knowledge, this is the first systematic review to directly compare estimates of the rate of opioid-related mortality between buprenorphine and methadone.

## 2. Methods

We conducted this systematic review in accordance with a pre-specified protocol (PROSPERO number: CRD42021234769), and we reported our findings following the Preferred Reporting Items for Systematic Reviews and Meta-Analyses (PRISMA) 2020 checklist [33].

### 2.1. Search Strategy

We conducted a systematic literature search of studies comparing buprenorphine and methadone among people with opioid use disorder from 1978 to February 6, 2021 in Medline, Embase, PsycINFO, and Web of Science databases. We also scanned the bibliographies of the included articles for additional references. We tailored our search strategy to each database, and search terms and keywords included those related to buprenorphine, methadone, opioid use disorder, and opioid-related mortality (see **Supplementary Materials** for Search Strategy). We restricted the search period to 1978 and onward because buprenorphine was first identified as a treatment option for treating addiction in 1978 [34]. There were no restrictions on the language of publication, and the search was updated on April 8, 2023.

### 2.2. Inclusion and Exclusion Criteria

We included observational studies (e.g., cohort or case-control studies) whose objective was to compare the risk of opioid-related mortality from buprenorphine with that from methadone in populations undergoing medication-assisted treatment for opioid use disorder. We excluded randomized controlled trials because they often have shorter follow-up period, resulting in an insufficient number of mortality cases to draw inferences [35]. We also excluded cross-sectional studies, letters to the editor, commentaries, editorials, and conference abstracts, as these studies do not contain the information needed to adequately assess their methodological quality.

### 2.3. Outcome Measures

The primary outcome of interest was drug-related mortality, including poisoning and fatal overdose events.

### 2.4. Study Selection

Two independent reviewers (JL and SO) screened titles and abstracts, and then conducted a full-text review of all articles retrieved from the databases for eligibility (study selection) based on specified inclusion/exclusion criteria. Disagreements between the two reviewers were resolved through consensus or adjudication by a third independent reviewer (DP).

### 2.5. Data Extraction

Two independent reviewers (JL and IF) extracted data using a pre-piloted, standardized data extraction form (see **Supplementary Materials**), with disagreements resolved through consensus or adjudication by a third independent reviewer (DP). Descriptive texts and quantitative data extracted included study characteristics, baseline participant characteristics, and outcome data. Study characteristics included study design, setting, data source, duration of follow-up, opioid use disorder assessment method, inclusion/exclusion criteria, and sample size. Baseline participant characteristics included distribution of age, sex, and history of psychiatric disorders (if applicable). Finally, the outcome data included a measure of association (e.g., incidence rate ratios [IRR], odds ratios [OR], or hazard ratios [HR]) and 95% confidence interval (CI).

### 2.6. Assessment of Risk of Bias

Two reviewers (JL and IF) independently assessed the risk of bias for all included studies. For these studies, we used the Risk of Bias in Non-Randomized Studies - of Interventions (ROBINS-I) tool to examine potential biases in seven domains [36]. These include (1) Confounding, (2) Selection bias, (3) Misclassification of intervention groups, (4) Deviations from intended intervention, (5) Missing data, (6) Outcome measurement error, and (7) Selective reporting. Based on the assessment of each domain, we assigned the overall risk of bias as low, moderate, serious, or critical, where the overall risk was determined by the highest risk assigned in any individual domain. In addition, we assessed the presence of other forms of biases that stem from inappropriate study designs in pharmacoepidemiology, including prevalent user bias (i.e., inability to capture early events due to prevalent users having survived initial stages of medication use), informative censoring (i.e., stopping follow-up at an arbitrarily set calendar date), and left truncation (i.e., initiation of follow-up at an arbitrary calendar date).

## 3. Results

### 3.1. Search Results

The database searches identified 3109 publications (**Figure 1**). We removed 1330 duplicates and an additional 1697 studies upon review of the title and abstract. These studies were excluded because they were irrelevant to the research question or had non-observational study designs. Two reviewers independently examined the remaining 82 articles and excluded 75 of them (see **Supplementary Materials** for the list of articles for full-text review and reasons for exclusion). Our review included seven studies [37–43]. Since the number of studies was not sufficiently large for quantitative data synthesis, we conducted a systematic review without meta-analysis.

**Figure 1.**
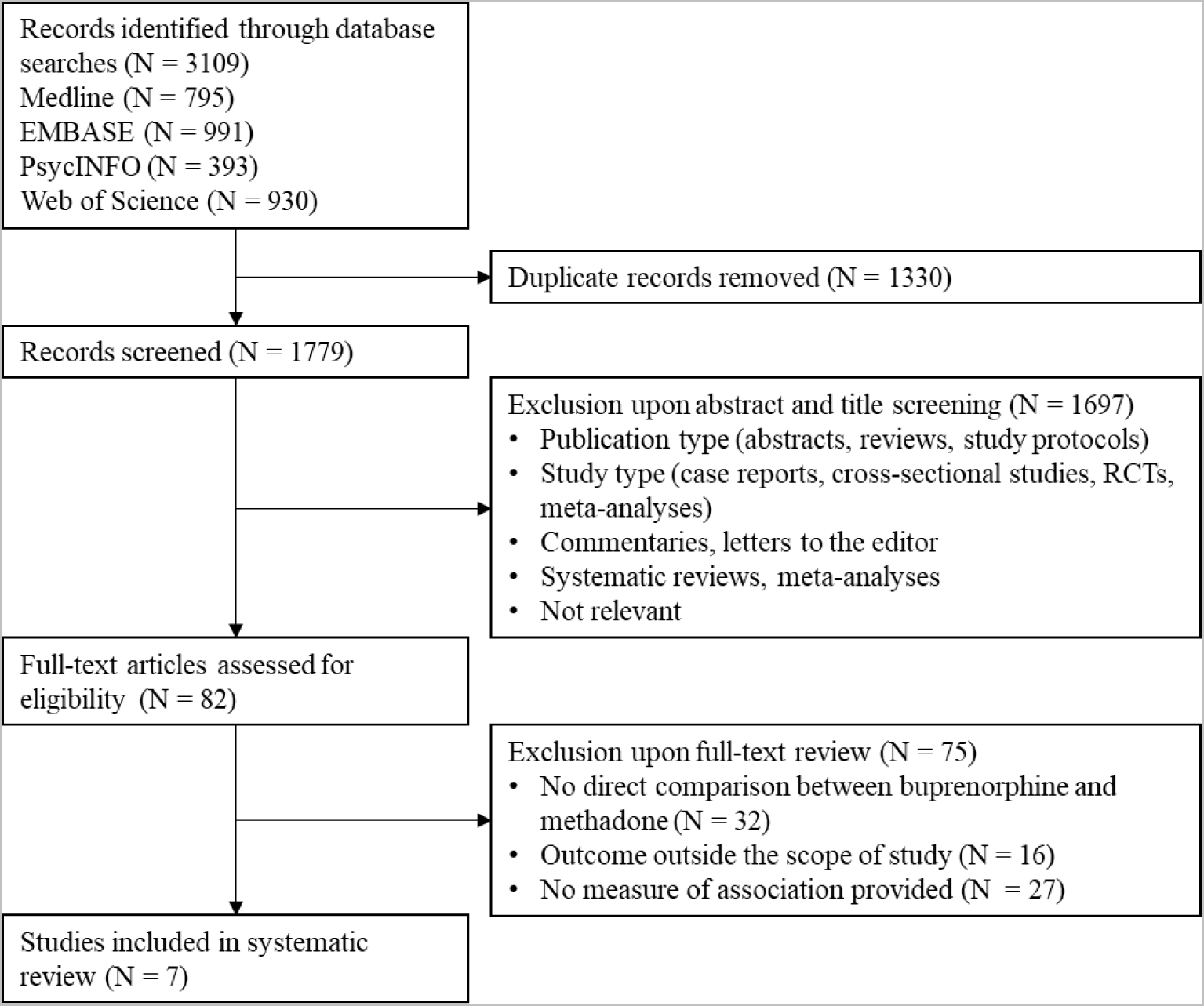
Selection of studies for systematic review

### 3.2. Study Characteristics

**Table 1** summarizes the characteristics of the six studies, which were conducted in Norway (N = 1) [37], the United Kingdom (UK) (N = 2) [38, 39], and Australia (N = 4) [40–43]. Four studies were retrospective cohort studies with a new-user design that included all patients who initiated OAT medications [38, 40, 42, 43], and three were administrative database studies that included only the OAT patients who died from exposure to either buprenorphine or methadone [37, 39, 41] (see **Table 1** for the exact study design terminologies presented by the authors of the included studies). All studies defined exposure to buprenorphine or methadone as treatment for OUD, and they excluded buprenorphine or methadone that was indicated as analgesics or cough suppressants. Study cohort size ranged from 200 to 45664 patients. Drug-related mortality rates associated with each drug were calculated based on the number of deaths per 1,000 persons, the number of deaths per person-years of drug exposure, or the number of drug prescriptions. Five studies reported the number of deaths and person-years associated with the two medications [37, 38, 40, 42, 43].

**Table 1.** Characteristics of observational studies comparing the risk of drug-related mortality associated with buprenorphine and methadone

| Study | Country | Data Source | Study Period | Design | N |
| --- | --- | --- | --- | --- | --- |
| Bech et al. (2019) [37] | Norway | (a) Norwegian Cause of Death Registry<br>(b) Norwegian Patient Registry | 01/2014-12/2015 | Observational register study<sup>#</sup> | 200 |
| Bell et al. (2009) [41] | Australia | Pharmaceutical Drugs of Addiction System | 04/2006-12/2006 | Data linkage study<sup>#</sup> | 16434 |
| Hickman et al. (2018) [38] | UK | (a) Clinical Practice Research Datalink<br>(b) Office for National Statistics (mortality data) | 1998-2014 | Retrospective cohort study | 5935 |
| Jones et al. (2022) [42] | Australia | Electronic Reporting and Recording of Controlled Drugs (formerly Pharmaceutical Drugs of Addiction System) | 2002-2017 | Retrospective cohort study | 45664 |
| Kimber et al. (2015) [40] | Australia | Pharmaceutical Drugs of Addiction System | 2001-2010 | Retrospective cohort study | 32033 |
| Larney et al. (2023) [43] | Australia | Electronic Reporting and Recording of Controlled Drugs (formerly Pharmaceutical Drugs of Addiction System) | 2002-2017 | Retrospective cohort study | 37764 |
| Marteau et al. (2015) [39] | England and Wales | (a) National Health Service<br>(b) Office for National Statistics ‘Deaths Related to Drug Poisoning in England and Wales’ | 2007-2012 | Retrospective administrative data study<sup>#</sup> | NI<sup>\$</sup> |

Abbreviations: OUD: Opioid use disorder; BUP: Buprenorphine; BNX: Buprenorphine-naloxone; MET: Methadone; OAT: Opioid agonist treatment; NI: No information
<sup>#</sup> The study design terminologies were those presented by the authors of the included studies.
<sup>\$</sup> Information on the number of patients was not available in the study. In addition, the rates of drug-related mortality for buprenorphine and methadone were calculated by using the number of prescriptions as the denominator. The authors of this study took the rate of mortality associated with methadone and divided by the rate of mortality associated with buprenorphine.

### 3.3. Risk of Drug-Related Poisoning Deaths

**Table 2** reports the effect estimates of the risk of drug-related mortality associated with buprenorphine and methadone. Three studies reported no difference between buprenorphine and methadone or only weak evidence in favour of buprenorphine [37, 42, 43]. The other four studies, on the other hand, reported a more favourable safety profile for buprenorphine (rate ratio ranging from 0.08 [95% CI = 0.01-0.48] in favour of buprenorphine to 6.23 [95% CI = 4.79- 8.10] against methadone) [38–41]. However, three studies restricted the study to only the patients whose cause of death involved buprenorphine or methadone, which may bias the findings and undermine the validity of results (see section on Risk of Bias for more detail) [37, 39, 41]. Therefore, we summarize below findings from four retrospective cohort studies with a new-user design [38, 40, 42, 43].

**Table 2.**
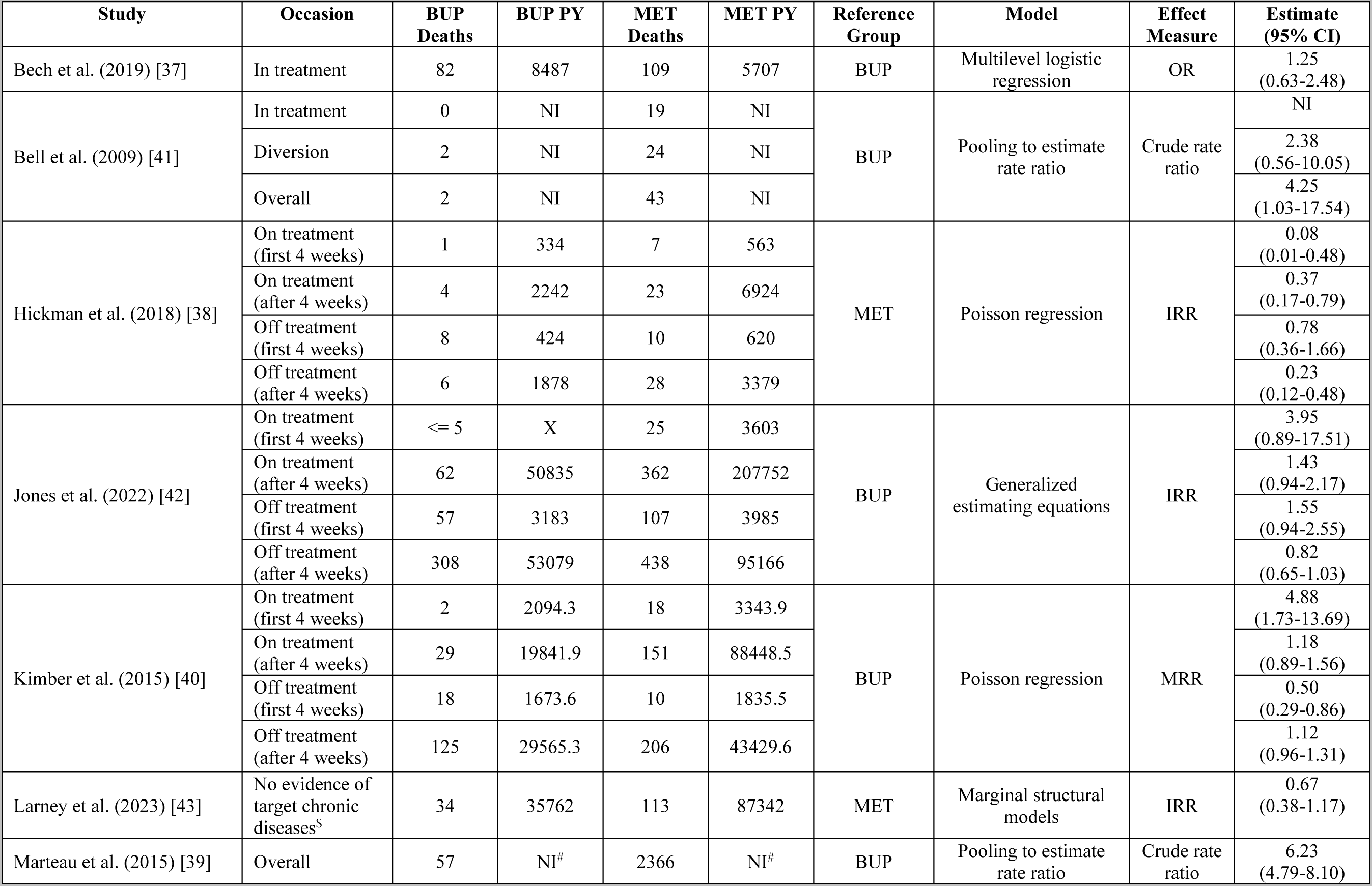
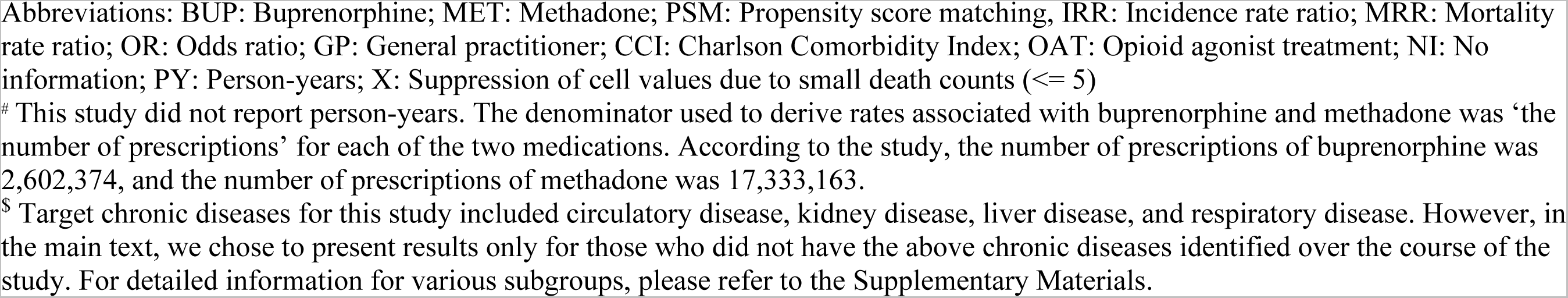
Effect estimates of the risk of drug-related mortality associated with buprenorphine and methadone.

| Study | Occasion | BUP Deaths | BUP PY | MET Deaths | MET PY | Reference Group | Model | Effect Measure | Estimate (95% CI) |
| --- | --- | --- | --- | --- | --- | --- | --- | --- | --- |
| Bech et al. (2019) [37] | In treatment | 82 | 8487 | 109 | 5707 | BUP | Multilevel logistic regression | OR | 1.25 (0.63-2.48) |
| Bell et al. (2009) [41] | In treatment | 0 | NI | 19 | NI | BUP | Pooling to estimate rate ratio | Crude rate ratio | NI |
|  | Diversion | 2 | NI | 24 | NI |  |  |  | 2.38 (0.56-10.05) |
|  | Overall | 2 | NI | 43 | NI |  |  |  | 4.25 (1.03-17.54) |
| Hickman et al. (2018) [38] | On treatment (first 4 weeks) | 1 | 334 | 7 | 563 | MET | Poisson regression | IRR | 0.08 (0.01-0.48) |
|  | On treatment (after 4 weeks) | 4 | 2242 | 23 | 6924 |  |  |  | 0.37 (0.17-0.79) |
|  | Off treatment (first 4 weeks) | 8 | 424 | 10 | 620 |  |  |  | 0.78 (0.36-1.66) |
|  | Off treatment (after 4 weeks) | 6 | 1878 | 28 | 3379 |  |  |  | 0.23 (0.12-0.48) |
| Jones et al. (2022) [42] | On treatment (first 4 weeks) | <= 5 | X | 25 | 3603 | BUP | Generalized estimating equations | IRR | 3.95 (0.89-17.51) |
|  | On treatment (after 4 weeks) | 62 | 50835 | 362 | 207752 |  |  |  | 1.43 (0.94-2.17) |
|  | Off treatment (first 4 weeks) | 57 | 3183 | 107 | 3985 |  |  |  | 1.55 (0.94-2.55) |
|  | Off treatment (after 4 weeks) | 308 | 53079 | 438 | 95166 |  |  |  | 0.82 (0.65-1.03) |
| Kimber et al. (2015) [40] | On treatment (first 4 weeks) | 2 | 2094.3 | 18 | 3343.9 | BUP | Poisson regression | MRR | 4.88 (1.73-13.69) |
|  | On treatment (after 4 weeks) | 29 | 19841.9 | 151 | 88448.5 |  |  |  | 1.18 (0.89-1.56) |
|  | Off treatment (first 4 weeks) | 18 | 1673.6 | 10 | 1835.5 |  |  |  | 0.50 (0.29-0.86) |
|  | Off treatment (after 4 weeks) | 125 | 29565.3 | 206 | 43429.6 |  |  |  | 1.12 (0.96-1.31) |
| Larney et al. (2023) [43] | No evidence of target chronic diseases<sup>s</sup> | 34 | 35762 | 113 | 87342 | MET | Marginal structural models | IRR | 0.67 (0.38-1.17) |
| Marteau et al. (2015) [39] | Overall | 57 | NI<sup>#</sup> | 2366 | NI<sup>#</sup> | BUP | Pooling to estimate rate ratio | Crude rate ratio | 6.23 (4.79-8.10) |

Abbreviations: BUP: Buprenorphine; MET: Methadone; PSM: Propensity score matching, IRR: Incidence rate ratio; MRR: Mortality rate ratio; OR: Odds ratio; GP: General practitioner; CCI: Charlson Comorbidity Index; OAT: Opioid agonist treatment; NI: No information; PY: Person-years; X: Suppression of cell values due to small death counts ( $\leq 5$ )
<sup>#</sup> This study did not report person-years. The denominator used to derive rates associated with buprenorphine and methadone was ‘the number of prescriptions’ for each of the two medications. According to the study, the number of prescriptions of buprenorphine was 2,602,374, and the number of prescriptions of methadone was 17,333,163.
<sup>\$</sup> Target chronic diseases for this study included circulatory disease, kidney disease, liver disease, and respiratory disease. However, in the main text, we chose to present results only for those who did not have the above chronic diseases identified over the course of the study. For detailed information for various subgroups, please refer to the Supplementary Materials.

Of these studies with low risk of bias in selection of participants in the study, the estimated risks of drug-related poisoning deaths were heterogeneous across time. A UK-based study by Hickman et al. (2018) showed that being on buprenorphine treatment was associated with 92% lower rate of death compared to being on methadone treatment when the OAT patients were ‘on’ treatment (i.e., having active prescription of OAT medications) during the first four weeks (IRR = 0.08; 95% CI = 0.01-0.48) [38]. Similarly, an Australian study by Kimber et al. (2015) showed that methadone was associated with an almost 5-fold increased rate of fatal overdose compared to buprenorphine during the same time window (IRR = 4.88; 95% CI = 1.73-13.69) [40]. A more recent Australian study by Jones et al. (2022) showed that methadone was associated with nearly four times the higher rate of fatal overdose compared to buprenorphine during the first four weeks of being ‘on’ treatment, although the results were not statistically significant (IRR = 3.95; 95% CI = 0.89-17.51) [42]. However, after the four-week treatment initiation period, the rate of drug-related mortality while ‘on’ OAT differed across the three studies. The UK study above reported that being on buprenorphine treatment continued to have a protective effect against drug-related poisoning deaths compared to being on methadone treatment (IRR = 0.37; 95% CI = 0.17-0.79) [38]. On the other hand, the two aforesaid Australian studies showed that the effect of the two medications on drug-related poisoning deaths did not differ, with IRR = 1.18 (95% CI = 0.89-1.56) and IRR = 1.43 (95% CI = 0.94-2.17) observed by Kimber et al. (2015) and Jones et al. (2022), respectively [40, 42]. Relatedly, another Australian study by Larney et al. (2023) demonstrated that the risk of opioid overdose mortality was lower while being on buprenorphine treatment than on methadone treatment, although the results were not statistically significant (IRR = 0.67; 95% CI = 0.38-1.17) [43]. The authors of this study also observed similar results for patients across different age groups as well as patients with kidney disease and liver disease (see **Supplementary Materials** for measures of association done by subgroups).

The rates of drug-related mortality between buprenorphine and methadone varied across treatment status as well. When the OAT patients were ‘off’ treatment (i.e., discontinued or no longer on buprenorphine or methadone), the UK study showed no significant differences in the rate of death following buprenorphine discontinuation compared to that following methadone discontinuation during the first four weeks (IRR = 0.78; 95% CI = 0.36-1.66) and lower rate of death in favour of buprenorphine after the initial four-week period (IRR = 0.23; 95% CI = 0.12-0.48) [38]. However, the Australian study by Kimber et al. (2015) concluded that the rate of death following methadone discontinuation was lower than that following buprenorphine discontinuation (IRR = 0.50; 95% CI = 0.29-0.86) during the first four weeks and no difference after the initial four-week period (IRR = 1.12; 95% CI = 0.96-1.32) [40]. In addition, the Australian study by Jones et al. (2022) revealed no difference in the rate of fatal overdose when ‘off’ treatment both during the first four weeks (IRR = 1.55; 95% CI = 0.94-2.55) and after the initial four-week period (IRR = 0.82; 95% CI = 0.65-1.03) [42].

### 3.4. Risk of Bias

We conducted the risk of bias assessment for each study using the ROBINS-I (**Table 3**; see **Supplementary Materials** for a more detailed assessment of risk of bias). Out of the seven studies identified, three studies were assigned a moderate risk of bias [38, 42, 43], two were assigned a serious risk of bias [37, 40], and two were assigned a critical risk of bias [39, 41]. One domain that led to an increase in the risk of bias was the ‘bias due to confounding’, which resulted from residual confounding due to failure to adjust for important confounders [37, 39–41]. One study used fractional polynomial regression models to assess the presence of residual confounding and then the rule-out method to establish how strong the residual confounding would have to be to explain the effect sizes [40]. The authors of this study concluded that the association is unlikely to have been influenced by residual confounding, but they did not adjust for many key confounders (e.g., history of overdose or psychiatric morbidities) nor take into consideration potential interactions between them. Another study used a propensity score method with inverse probability weighting to control for confounding [38], but the potential for unmeasured confounding could not be excluded. Two studies also attempted to adjust for time-varying confounding (i.e., covariates whose values change over the course of follow-up) through regression adjustment [38, 42]. Of note, these covariates could be in the causal pathway; thus, adjusting for them possibly introduced over-adjustment bias [44]. Other, more appropriate techniques to control for time-varying confounding such as marginal structural Cox proportional hazards models were not considered [45], except in Larney et al. (2023) [43].

**Table 3.** Bias assessment of studies directly comparing drug-related poisoning mortality risk associated with buprenorphine and methadone.

|  | **Studies** | Bech et al. (2019) [37] | Bell et al. (2009) [41] | Hickman et al. (2018) [38] | Jones et al. (2022) [42] | Kimber et al. (2015) [40] | Larney et al. (2023) [43] | Marteau et al. (2015) [39] |
| --- | --- | --- | --- | --- | --- | --- | --- | --- |
|  | **Domains** |  |  |  |  |  |  |  |
| **ROBINS-I** | **Confounding** | Serious | Critical | Moderate | Moderate | Serious | Moderate | Critical |
|  | **Selection of participants in the study** | Serious | Serious | Low | Low | Low | Low | Serious |
|  | **Classification of interventions** | Serious | Serious | Moderate | Moderate | Moderate | Moderate | Serious |
|  | **Deviations from intended intervention** | Low | Low | Low | Low | Low | Low | Low |
|  | **Missing data** | Low | Low | Moderate | Low | Low | Low | Low |
|  | **Measurement of outcomes** | Low | Low | Low | Low | Low | Low | Low |
|  | **Selection of the reported result** | Moderate | Moderate | Moderate | Low | Moderate | Low | Moderate |
|  | **Overall risk of bias** | Serious | Critical | Moderate | Moderate | Serious | Moderate | Critical |
| **Other Sources of Bias** | **Prevalent user bias** | X | X |  |  |  |  | X |
|  | **Informative censoring** |  |  | X |  |  |  |  |
|  | **Left truncation** | X | X |  |  |  |  | X |

Another domain that led to an increase in the risk of bias was the ‘bias in selection of participants in the study’. Two studies included only the patients who experienced drug-related poisoning death and then retrospectively ascertained exposure to buprenorphine or methadone [37, 41]. These studies failed to account for person-time accumulated by the patients who were prescribed OAT for OUD but did not experience overdose death over the course of follow-up. In addition, due to left truncation and associated challenges in identifying OAT initiation, the possibility of prevalent user bias could not be excluded for these database studies. One study included all cases of drug-related poisoning death, including the patients who have not been prescribed OAT medications [39]. The mortality cases include those that involved buprenorphine or methadone from outside the healthcare system as well as deceased individuals who never utilized the healthcare system. Inclusion of such cases biases the relationship between drug-related poisoning mortality and medications prescribed as OAT within the healthcare system. Four retrospective cohort studies were new-user studies that followed OAT patients until overdose death or censoring, which mitigated the risk of bias in selection of participants into the study [38, 40, 42, 43]. However, one study censored patients after 12 months of OAT discontinuation [38]. According to the authors, this was done “to exclude cumulative dilution of mortality risks”, but it may have excluded treatment re-initiation or deaths that occur during the ‘off’ treatment period. Therefore, selection bias due to informative censoring is possible.

All identified studies defined exposure to buprenorphine or methadone based on records from administrative databases. Four studies used the time-varying exposure definition with grace periods to control for potential exposure misclassification [38, 40, 42, 43], but the addition of residual effect period does not completely eliminate misclassification of exposure time window. Hence, these studies were judged to be at moderate risk of bias in classification of interventions. Three studies did not identify with which of buprenorphine or methadone the patients initiated the OAT or how long they have stayed in the OAT prior to study entry [37, 39, 41]. Among these three, one study also classified whether mortality occurred during or after discontinuation of OAT (i.e., diversion) using database records of drug prescription [41], which further elevated the risk of exposure misclassification. Therefore, the above three studies were judged to be at severe risk of bias in classification of interventions. Finally, five studies were ascribed a moderate risk of bias in ‘selection of reported results’ due to the absence of a pre-specified protocol [37–41], and two were ascribed low risk due availability of a pre-specified protocol and analysis consistent with it [42, 43].

## 4. Discussion

The objective of our systematic review was to synthesize available evidence on the comparative safety of buprenorphine and methadone and to assess the methodological strengths and limitations of this literature. We identified seven observational studies, with variations with respect to the types of databases used and study design characteristics, that met our inclusion criteria. These studies came from three different countries (Norway, UK, and Australia), each with different clinical practice guidelines and regulatory environment that dictates OAT prescribing patterns. We found mixed evidence of comparative mortality risk associated with buprenorphine and methadone. Using ROBINS-I to assess the overall study quality, we determined that 3 study had a moderate risk of bias, 2 had a serious risk, and 2 had a critical risk. The most serious risks underlying these studies were residual confounding and selection bias in the form of prevalent user bias, informative censoring, and left truncation. We did not conduct meta-analysis for this review because there were only three studies with moderate risk of bias based on our bias assessment using the ROBINS-I tool, and two of the studies came from the same data source with an overlapping time window [42, 43].

For estimating long-term OAT risks including mortality, observational studies have been widely accepted as the best evidence for clinical and policy decision-making, since randomized controlled trials, which often face budgetary constraints, have low statistical power with a shorter follow-up period [35]. However, several methodological shortcomings in the observational studies included in this systematic review have important implications for public health stakeholders and patients around the world. For example, in Canada, buprenorphine is currently recommended as the first-line therapy over methadone due to the former’s “superior safety profile” [13, 46]. However, some of the evidence of mortality risk underlying these guidelines have come from observational studies with serious methodological flaws, including residual confounding, selection bias, prevalent user bias, and left truncation [39, 41]. Further, previous systematic reviews that concluded superiority of buprenorphine to methadone in mitigating drug-related poisoning mortality were based on indirect comparisons (i.e., calculation of mortality rates from ‘on’ vs. ‘off’ OAT for each medication separately) as opposed to direct comparisons [6, 32]. Therefore, additional head-to-head comparisons of buprenorphine and methadone in observational settings are needed to better understand comparative safety and effectiveness of these two medications in real-world clinical practice.

For more methodologically robust observational evidence, cohort studies would need to be restricted to “new users” of OAT medications (i.e., patients who initiate buprenorphine or methadone for the first time). The rationale is that in studies with prevalent users, covariates for the patients at study cohort entry may be affected by the drug itself, resulting in improper adjustment of confounders that were measured after treatment initiation [47]. In addition, entry into the cohort should ideally take place when the individuals initiate treatment as opposed to a fixed calendar time in order to minimize bias from left truncation [48]. With evidence of OAT re-initiation after discontinuation for as long as 18 months [49], follow-up should occur until drug-related mortality, death from other causes, departure from the database, or end of the study period in order to capture deaths and person-time that occur during the OAT discontinuation period and prevent informative censoring [50]. Relatedly, longer time windows for follow-up should be considered for studies examining mortality risk associated with OAT medication use. Finally, to mitigate residual confounding, more potential confounders should be included in the analyses, and in the presence of time-varying confounders, marginal structural models should be considered.

Our study has several strengths. First, the study followed a pre-specified protocol, with the comprehensive systematic search of four databases. Second, to assess the methodological quality of the included studies, we used ROBINS-I, a state-of-the-art tool that enables a thorough assessment of the risk of bias in several important domains, including confounding and selection bias. Finally, we also examined the presence of three additional sources of bias prevalent in pharmacoepidemiologic studies, which supplemented ROBINS-I and ensured a comprehensive assessment of the methodological quality of the included studies.

Our study also has a few limitations. First, our review is affected by the methodological limitations of the included studies, such as residual confounding arising from clinical and sociodemographic data not captured by administrative databases. As a result of these limitations, there were only three studies with a moderate risk of bias from which we could draw substantive conclusions. Second, due to heterogeneity in study design, study definitions, and study populations in addition to the modest quality of this literature, we were unable to pool results across studies. An important source of between-study heterogeneity includes the rate ratios presented by the studies in this review, which were derived from different denominators (e.g., per 1,000 people; person-years; and number of prescriptions). Third, evidence that comprise our systematic review comes from several different countries, each with its own regulatory environments and clinical practice guidelines that dictate prescription of OAT [51]. Even if we were able to pool results across studies, the substantive differences in the delivery of care across different countries limit the generalizability of our study findings. Fourth, some of the studies included in this review have drawn their data from the same database with overlapping time windows [40–43]. Observations from these studies were unlikely to be independent, further highlighting the lack of independent, high-quality studies in this literature. However, through the comprehensive assessment of the methodological quality of this literature, we were able to reveal the need for a greater number of methodologically rigorous observational studies to better understand comparative risks of drug-related mortality associated with buprenorphine and methadone.

## 5. Conclusion

Although several studies have conducted a direct head-to-head comparison of buprenorphine and methadone in relation to drug-related mortality, interpretation of these results has been difficult due to the methodological limitations of this literature. The three highest quality studies in the literature, which were conducted in the UK and Australia, have shown discrepancies in their findings, which underscore uncertainties in the comparative effectiveness surrounding buprenorphine and methadone. The UK study suggests that buprenorphine has a safer profile than methadone in mitigating the risk of drug-related mortality, whereas the Australian studies suggest that the rates of fatal overdose associated with the two medications are not different. Due to limited evidence in the literature and heterogeneity in clinical practice and regulatory environment in the studies that were included in our review, generalizability of the findings for our research question remains difficult at this time. Therefore, to provide a more accurate long-term comparative safety profile for these two medications, additional observational studies with large sample sizes and methodological rigour are warranted.

## Data Availability

All data produced in the present work are contained in the manuscript.

## Acknowledgements

None

## Contributions

Jihoon Lim contributed to study conception, performed literature search, conducted data extraction, assessed the risk of bias in included studies, and drafted the manuscript. Imen Farhat conducted data extraction, assessed the risk of bias in included studies, and reviewed the manuscript for important intellectual content. Dr. Antonios Douros provided methodological expertise for several of the criteria used in the risk of bias assessment and reviewed the manuscript for important intellectual content. Soukaina Ouizzane performed literature search.

Dr. Dimitra Panagiotoglou supervised the project, contributed to study conception, and reviewed the manuscript for important intellectual content. All authors read, contributed to, and approved the final manuscript.

## Registration

This systematic review has been registered with PROSPERO (https://www.crd.york.ac.uk/prospero) (identifier CRD42021234769).

---

PRISMA 2020 Checklist.

PRISMA 2020 Checklist
| Section and Topic | Item # | Checklist item | Location where item is reported |
| --- | --- | --- | --- |
| **TITLE** |  |  |  |
| Title | 1 | Identify the report as a systematic review. | Title Page |
| **ABSTRACT** |  |  |  |
| Abstract | 2 | See the PRISMA 2020 for Abstracts checklist. | Abstract Page |
| **INTRODUCTION** |  |  |  |
| Rationale | 3 | Describe the rationale for the review in the context of existing knowledge. | Section 1 |
| Objectives | 4 | Provide an explicit statement of the objective(s) or question(s) the review addresses. | Section 1 |
| **METHODS** |  |  |  |
| Eligibility criteria | 5 | Specify the inclusion and exclusion criteria for the review and how studies were grouped for the syntheses. | Section 2.2 |
| Information sources | 6 | Specify all databases, registers, websites, organisations, reference lists and other sources searched or consulted to identify studies. Specify the date when each source was last searched or consulted. | Section 2.1 |
| Search strategy | 7 | Present the full search strategies for all databases, registers and websites, including any filters and limits used. | Supplementary Materials |
| Selection process | 8 | Specify the methods used to decide whether a study met the inclusion criteria of the review, including how many reviewers screened each record and each report retrieved, whether they worked independently, and if applicable, details of automation tools used in the process. | Section 2.4 |
| Data collection process | 9 | Specify the methods used to collect data from reports, including how many reviewers collected data from each report, whether they worked independently, any processes for obtaining or confirming data from study investigators, and if applicable, details of automation tools used in the process. | Section 2.5 |
| Data items | 10a | List and define all outcomes for which data were sought. Specify whether all results that were compatible with each outcome domain in each study were sought (e.g. for all measures, time points, analyses), and if not, the methods used to decide which results to collect. | Section 2.3 Protocol |
|  | 10b | List and define all other variables for which data were sought (e.g. participant and intervention characteristics, funding sources). Describe any assumptions made about any missing or unclear information. | Tables 1 & 2 Supplementary Materials |
| Study risk of bias assessment | 11 | Specify the methods used to assess risk of bias in the included studies, including details of the tool(s) used, how many reviewers assessed each study and whether they worked independently, and if applicable, details of automation tools used in the process. | Section 2.6 |
| Effect measures | 12 | Specify for each outcome the effect measure(s) (e.g. risk ratio, mean difference) used in the synthesis or presentation of results. | Section 2.5 |
| Synthesis methods | 13a | Describe the processes used to decide which studies were eligible for each synthesis (e.g. tabulating the study intervention characteristics and comparing against the planned groups for each synthesis (item #5)). | NA – Not a meta-analysis |
|  | 13b | Describe any methods required to prepare the data for presentation or synthesis, such as handling of missing summary statistics, or data conversions. | NA – Not a meta-analysis |
|  | 13c | Describe any methods used to tabulate or visually display results of individual studies and syntheses. | NA – Not a meta-analysis |
|  | 13d | Describe any methods used to synthesize results and provide a rationale for the choice(s). If meta-analysis was performed, describe the model(s), method(s) to identify the presence and extent of statistical heterogeneity, and software package(s) used. | NA – Not a meta-analysis |
|  | 13e | Describe any methods used to explore possible causes of heterogeneity among study results (e.g. subgroup analysis, meta-regression). | NA – Not a meta- |

PRISMA 2020 Checklist
| Section and Topic | Item # | Checklist item | Location where item is reported |
| --- | --- | --- | --- |
|  |  |  | analysis |
|  | 13f | Describe any sensitivity analyses conducted to assess robustness of the synthesized results. | NA – Not a meta-analysis |
| Reporting bias assessment | 14 | Describe any methods used to assess risk of bias due to missing results in a synthesis (arising from reporting biases). | NA – Not a meta-analysis |
| Certainty assessment | 15 | Describe any methods used to assess certainty (or confidence) in the body of evidence for an outcome. | NA – Not a meta-analysis |
| **RESULTS** |  |  |  |
| Study selection | 16a | Describe the results of the search and selection process, from the number of records identified in the search to the number of studies included in the review, ideally using a flow diagram. | Section 3.1 |
|  | 16b | Cite studies that might appear to meet the inclusion criteria, but which were excluded, and explain why they were excluded. | Supplementary Materials |
| Study characteristics | 17 | Cite each included study and present its characteristics. | Section 3.2<br>Table 1 |
| Risk of bias in studies | 18 | Present assessments of risk of bias for each included study. | Section 3.4<br>Table 3 |
| Results of individual studies | 19 | For all outcomes, present, for each study: (a) summary statistics for each group (where appropriate) and (b) an effect estimate and its precision (e.g. confidence/credible interval), ideally using structured tables or plots. | Section 3.3<br>Table 2 |
| Results of syntheses | 20a | For each synthesis, briefly summarise the characteristics and risk of bias among contributing studies. | NA – Not a meta-analysis |
|  | 20b | Present results of all statistical syntheses conducted. If meta-analysis was done, present for each the summary estimate and its precision (e.g. confidence/credible interval) and measures of statistical heterogeneity. If comparing groups, describe the direction of the effect. | NA – Not a meta-analysis |
|  | 20c | Present results of all investigations of possible causes of heterogeneity among study results. | NA – Not a meta-analysis |
|  | 20d | Present results of all sensitivity analyses conducted to assess the robustness of the synthesized results. | NA – Not a meta-analysis |
| Reporting biases | 21 | Present assessments of risk of bias due to missing results (arising from reporting biases) for each synthesis assessed. | NA – Not a meta-analysis |
| Certainty of evidence | 22 | Present assessments of certainty (or confidence) in the body of evidence for each outcome assessed. | NA – Not a meta-analysis |
| **DISCUSSION** |  |  |  |
| Discussion | 23a | Provide a general interpretation of the results in the context of other evidence. | Section 4 |
|  | 23b | Discuss any limitations of the evidence included in the review. | Section 4 |
|  | 23c | Discuss any limitations of the review processes used. | Section 4 |

PRISMA 2020 Checklist
| Section and Topic | Item # | Checklist item | Location where item is reported |
| --- | --- | --- | --- |
|  | 23d | Discuss implications of the results for practice, policy, and future research. | Sections 4 & 5 |
| **OTHER INFORMATION** |  |  |  |
| Registration and protocol | 24a | Provide registration information for the review, including register name and registration number, or state that the review was not registered. | P. 16 |
|  | 24b | Indicate where the review protocol can be accessed, or state that a protocol was not prepared. | P. 16 |
|  | 24c | Describe and explain any amendments to information provided at registration or in the protocol. | See Protocol on PROSPERO |
| Support | 25 | Describe sources of financial or non-financial support for the review, and the role of the funders or sponsors in the review. | P. 16 |
| Competing interests | 26 | Declare any competing interests of review authors. | P. 16 |
| Availability of data, code and other materials | 27 | Report which of the following are publicly available and where they can be found: template data collection forms; data extracted from included studies; data used for all analyses; analytic code; any other materials used in the review. | Supplementary Materials |

From: Page MJ, McKenzie JE, Bossuyt PM, Boutron I, Hoffmann TC, Mulrow CD, et al. The PRISMA 2020 statement: an updated guideline for reporting systematic reviews. BMJ 2021;372:n71. doi: 10.1136/bmj.n71
For more information, visit: http://www.prisma-statement.org/

