## Supplemental Tables 1-5 for "Risk of opioid-related mortality associated with buprenorphine versus methadone: A systematic review of observational studies"

### SUPPLEMENTARY MATERIALS

**Title**: Relative effectiveness of medications for opioid-related disorders: A systematic review and network meta-analysis

**Authors**: Jihoon Lim MS, Imen Farhat MSc, Antonios Douros MD PhD, Soukaina Ouizzane BSc, Dimitra Panagiotoglou PhD

**Table S1**. Search strategy

| Database | Algorithm |
|---|---|
| MEDLINE | 1. exp opioid-related disorders/<br>2. ((drug OR substance OR opioid* OR opiat*) adj3 (disorder* OR addict* OR abuse* OR depend*)).ti,ab,kf.<br>3. 1 OR 2<br>4. exp buprenorphine/ OR buprenorphine.ti,ab,kf.<br>5. exp methadone/ OR methadone.ti,ab,kf.<br>6. 4 OR 5<br>7. ((opioid OR drug) adj2 (overdose* OR mortal* OR death* OR fatal* OR poison*)).ti,ab,kf.<br>8. 3 AND 6 AND 7<br>9. exp animals/ not humans.sh.<br>10. 8 NOT 9<br>11. limit 10 to yr="1978 -Current" |
| EMBASE | 1. exp addiction/<br>2. exp drug abuse/<br>3. ((drug OR substance OR opioid* OR opiat*) adj3 (disorder* OR addict* OR abuse* OR depend*)).ab,ti.<br>4. 1 OR 2 OR 3<br>5. exp buprenorphine/ OR buprenorphine.ab,ti.<br>6. exp methadone/ OR methadone.ab,ti.<br>7. 5 OR 6<br>8. ((opioid OR drug) adj2 (overdose* OR mortal* OR death* OR fatal* OR poison*)).ti,ab,kw,dj.<br>9. 4 AND 7 AND 8<br>10. limit 9 to (human AND embase AND yr="1978 -Current") |
| PsycINFO | 1. exp addiction/<br>2. exp drug abuse/<br>3. ((drug OR substance OR opioid* OR opiat*) adj3 (disorder* OR addict* OR abuse* OR depend*)).ti,ab,id.<br>4. 1 OR 2 OR 3<br>5. exp buprenorphine/ OR buprenorphine.ti,ab,id.<br>6. exp methadone/ OR methadone.ti,ab,id.<br>7. 5 OR 6<br>8. ((opioid OR drug) adj2 (overdose* OR mortal* OR death* OR fatal* OR poison*)).ti,ab,id.<br>9. 4 AND 7 AND 8<br>10. exp animals/ not humans.sh.<br>11. 9 NOT 10<br>12. Limit 11 to yr="1978 -Current" |
| Web of Science | 1. TS=(addiction)<br>2. TS=(drug abuse)<br>3. TS=((drug OR substance OR opioid* OR opiat*) NEAR/3 (disorder* OR addict* OR abuse* OR depend*))<br>4. 1 OR 2 OR 3<br>5. TS=(buprenorphine)<br>6. TS=(methadone)<br>7. 5 OR 6<br>8. TS=((opioid OR drug) NEAR/2 (overdose* OR mortal* OR death* OR fatal* OR poison*))<br>9. 4 AND 7 AND 8<br>10. TS=(animal* not (human* or patient*))<br>11. 9 NOT 10<br>12. PY=(1978-2021)<br>13. 11 AND 12 |

**Table S2**. Articles selected for full-text review and reasons for exclusion

| Last Name of First Author (Publication Year) | Article Title | Reason(s) for exclusion |
|---|---|---|
| Alderbergenov et al. (2022) | Methadone and buprenorphine-related deaths among people prescribed and not prescribed Opioid Agonist Therapy during the COVID-19 pandemic in England | No measure of association calculated |
| Alexandridis et al. (2019) | Associations between implementation of Project Lazarus and opioid analgesic dispensing and buprenorphine utilization in North Carolina, 2009-2014 | Outcome outside the scope of study |
| Ali et al. (2019) | Opioid Use Disorder and Prescribed Opioid Regimens: Evidence from Commercial and Medicaid Claims, 2005-2015 | Outcome outside the scope of study |
| Alinski et al. (2020) | Receipt of Addiction Treatment after Opioid Overdose among Medicaid-Enrolled Adolescents and Young Adults | Outcome outside the scope of study |
| Bagley et al. (2020) | Characteristics and Receipt of Medication Treatment Among Young Adults Who Experience a Nonfatal Opioid-Related Overdose | Outcome outside the scope of study |
| Bauer et al. (2008) | Mortality in opioid-maintained patients after release from an addiction clinic | No measure of association calculated |
| Bech et al. (2019) | Mortality and causes of death among patients with opioid use disorder receiving opioid agonist treatment: a national register study | Included |
| Bell et al. (2009) | Comparing overdose mortality associated with methadone and buprenorphine treatment | Included |
| Bhatraju et al. (2021) | Mortality in an opioid treatment program | No measure of association calculated |
| Buresh et al. (2021) | Treatment of opioid use disorder in primary care | No measure of association calculated |
| Burns et al. (2022) | Duration of medication treatment for opioid-use disorder and risk of overdose among Medicaid enrollees in 11 states: a retrospective cohort study | No direct comparison between buprenorphine and methadone |
| Caplehorn et al. (1999) | Mortality associated with New South Wales methadone programs in 1994: Lives lost and saved | No direct comparison between buprenorphine and methadone |
| Care et al. (2020) | Trends in severe opioid-related poisonings and fatalities reported to the Paris poison control center - a 10-year retrospective observational study | No measure of association calculated |
| Clausen et al. (2009) | Mortality among opiate users: opioid maintenance therapy, age and causes of death | No direct comparison between buprenorphine and methadone |
| Cornish et al. (2010) | Risk of death during and after opiate substitution treatment in primary care: prospective observational study in UK General Practice Research Database | No direct comparison between buprenorphine and methadone |
| Cousins et al. (2020) | Do interruptions to the continuity of methadone maintenance treatment (MMT) in specialist addiction settings increase the risk of drug-related poisoning (DRP) deaths? A retrospective cohort study | No direct comparison between buprenorphine and methadone |
| Cousins et al. (2011) | Risk of drug-related mortality during periods of transition in methadone maintenance treatment: A cohort study | No direct comparison between buprenorphine and methadone |
| Dasgupta et al. (2010) | Post-marketing Surveillance of Methadone and Buprenorphine in the United States | Outcome outside the scope of study |

| | | |
|---|---|---|
| Degenhardt et al. (2009) | Mortality among clients of a state-wide opioid pharmacotherapy program over 20 years: Risk factors and lives saved | No measure of association calculated |
| Dermengiu et al. (2013) | Drug related deaths between 2008 and 2011. A retrospective study in 32 Romanian counties | No measure of association calculated |
| Dermengiu et al. (2011) | Drugs of abuse identified in the National Institute of Legal Medicine Mina Minovici Bucharest 2010 | No measure of association calculated |
| Eastwood et al. (2017) | Effectiveness of treatment for opioid use disorder: A national, five-year, prospective, observational study in England | Outcome outside the scope of study |
| Faggiano et al. (2020) | Opioid overdose risk during and after drug treatment for heroin dependence: An incidence density case-control study nested in the vedette cohort | No direct comparison between buprenorphine and methadone |
| Fernandez-Calderon et al. (2017) | Drug-induced deaths in Southern Spain: profiles and associated characteristics | Outcome outside the scope of study |
| Fine et al. (2021) | Office-based addiction treatment retention and mortality among people experiencing homelessness | Outcome outside the scope of study |
| Fountain et al. (2019) | Deaths by poisoning in New Zealand, 2008-2013 | No measure of association calculated |
| Fugelstad et al. (2019) | Opioid-related deaths and previous care for drug use and pain relief in Sweden | No measure of association calculated |
| Fulton-Kehoe et al. (2015) | Opioid Poisonings in Washington State Medicaid: Trends, Dosing, and Guidelines | No measure of association calculated |
| Gibson et al. (2007) | Mortality related to pharmacotherapies for opioid dependence: a comparative analysis of coronial records | No direct comparison between buprenorphine and methadone |
| Glanz et al. (2022) | The association between buprenorphine treatment duration and mortality: a multi-site cohort study of people who discontinued treatment | No direct comparison between buprenorphine and methadone |
| Gomes et al. (2022) | Duration of use and outcomes among people with opioid use disorder initiating methadone and buprenorphine in Ontario: a population-based propensity-score matched cohort study | Outcome outside the scope of study |
| Gottlieb et al. (2022) | A comparison of mortality rates for buprenorphine versus methadone treatments for opioid use disorder | No measure of association provided |
| Hakkinen et al. (2012) | Comparison of fatal poisonings by prescription opioids | Outcome outside the scope of study |
| Hall et al. (2000) | Trends in opiate-related deaths in the United Kingdom and Australia, 1985-1995 | No measure of association calculated |
| Hall et al. (2008) | Patterns of abuse among unintentional pharmaceutical overdose fatalities | Outcome outside the scope of study |
| Hallowell et al. (2021) | History of Methadone and Buprenorphine Opioid Agonist Therapy Among People Who Died of an Accidental Opioid-Involved Overdose: Rhode Island, January 1, 2018–June 30, 2020 | No measure of association calculated |
| Hammersley et al. (1995) | Drugs associated with drug-related deaths in Edinburgh and Glasgow, November 1990 to October 1992 | No measure of association calculated |
| Handanagic et al. (2019) | Overdose mortality rates in Croatia and factors associated with self-reported drug overdose among persons who inject drugs in three Croatian cities | Outcome outside the scope of study |
| Handley et al. (2014) | Drugs and other chemicals involved in fatal poisoning in England and Wales during 2000-2011 | No measure of association calculated |
| Hickman et al. (2018) | The impact of buprenorphine and methadone on mortality: a primary care cohort study in the United Kingdom | Included |

| | | |
|---|---|---|
| Huang et al. (2013) | Factors associated with mortality among heroin users after seeking treatment with methadone: A population-based cohort study in Taiwan | No direct comparison between buprenorphine and methadone |
| Iwanicki et al. (2018) | Consistency Between Opioid-Related Mortality Trends Derived from Poison Center and National Vital Statistics System, United States, 2006-2016 | No measure of association calculated |
| Jones et al. (2022) | The impact of opioid agonist treatment on fatal and non-fatal drug overdose among people with a history of opioid dependence in NSW, Australia, 2001-2018: Findings from the OATS retrospective linkage study | Included |
| Jones et al. (2023) | Association of Receipt of Opioid Use Disorder-Related Telehealth Services and Medications for Opioid Use Disorder With Fatal Drug Overdoses Among Medicare Beneficiaries Before and During the COVID-19 Pandemic | No direct comparison between buprenorphine and methadone |
| Kalkman et al. (2019) | Trends in use and misuse of opioids in the Netherlands: a retrospective, multi-source database study | No direct comparison between buprenorphine and methadone |
| Karmali et al. (2020) | The role of substance use disorders in experiencing a repeat opioid overdose, and substance use treatment patterns among patients with a non-fatal opioid overdose | Outcome outside the scope of study |
| Kelty et al. (2017) | Fatal and non-fatal opioid overdose in opioid dependent patients treated with methadone, buprenorphine or implant naltrexone | No direct comparison between buprenorphine and methadone |
| Kelty et al. (2018) | Morbidity and mortality in opioid dependent patients after entering an opioid pharmacotherapy compared with a cohort of non-dependent controls | No direct comparison between buprenorphine and methadone |
| Kimber et al. (2015) | Mortality risk of opioid substitution therapy with methadone versus buprenorphine: A retrospective cohort study | Included |
| Krawczyk et al. (2020) | Opioid agonist treatment and fatal overdose risk in a state-wide US population receiving opioid use disorder services | No direct comparison between buprenorphine and methadone |
| Krawczyk et al. (2020) | Opioid agonist treatment is highly protective against overdose death among a U.S. statewide population of justice-involved adults | No direct comparison between buprenorphine and methadone |
| Larney et al. (2014) | Opioid substitution therapy as a strategy to reduce deaths in prison: retrospective cohort study | No direct comparison between buprenorphine and methadone |
| Larney et al. (2023) | Does opioid agonist treatment reduce overdose mortality risk in people who are older or have physical comorbidities? Cohort study using linked administrative health data in New South Wales, Australia, 2002–17 | Included |
| Larochelle et al. (2018) | Medication for Opioid Use Disorder After Nonfatal Opioid Overdose and Association with Mortality A Cohort Study | No direct comparison between buprenorphine and methadone |
| Lev et al. (2015) | Methadone related deaths compared to all prescription related deaths | No measure of association calculated |
| Lewer et al. (2020) | Life expectancy of people who are dependent on opioids: A cohort study in New South Wales, Australia | Outcome outside the scope of study |
| Lim et al. (2022) | Association between jail-based methadone or buprenorphine treatment for opioid use disorder and overdose mortality after release from New York City jails 2011-2017 | No direct comparison between buprenorphine and methadone |
| Lin et al. (2019) | Changing Trends in Opioid Overdose Deaths and Prescription Opioid Receipt Among Veterans | No direct comparison between buprenorphine and methadone |

| | | |
|---|---|---|
| Marsden et al. (2017) | Does exposure to opioid substitution treatment in prison reduce the risk of death after release? A national prospective observational study in England | No direct comparison between buprenorphine and methadone |
| Marteau et al. (2015) | The relative risk of fatal poisoning by methadone or buprenorphine within the wider population of England and Wales | Included |
| Megarbane et al. (2010) | Prospective comparative assessment of buprenorphine overdose with heroin and methadone: Clinical characteristics and response to antidotal treatment | No measure of association calculated |
| Mintz et al. (2022) | Associations between Stimulant Prescriptions and Drug-related Poisoning Risk among Persons Receiving Buprenorphine Treatment for Opioid Use Disorder | No direct comparison between buprenorphine and methadone |
| Morgan et al. (2020) | Comparison of Rates of Overdose and Hospitalization After Initiation of Medication for Opioid Use Disorder in the Inpatient vs Outpatient Setting | Outcome outside the scope of study |
| Moe et al. (2021) | Death after emergency department visits for opioid overdose in British Columbia: a retrospective cohort analysis | Outcome outside the scope of study |
| Pearce et al. (2020) | Opioid agonist treatment and risk of mortality during opioid overdose public health emergency: Population based retrospective cohort study | No direct comparison between buprenorphine and methadone |
| Pierce et al. (2016) | Impact of treatment for opioid dependence on fatal drug-related poisoning: a national cohort study in England | No direct comparison between buprenorphine and methadone |
| Qeadan et al. (2022) | Epidemiological trends in opioid-only and opioid/polysubstance-related death rates among American Indian/Alaska Native populations from 1999 to 2019: a retrospective longitudinal ecological study | No measure of association calculated |
| Rezza et al. (1992) | Estimating the trend of the epidemic of drug use in Italy, 1985-89 | No measure of association calculated |
| Shastry et al. (2022) | Prior use of medications for opioid use disorder in ED patients with opioid overdose: prevalence, misuse and overdose severity | No direct comparison between buprenorphine and methadone |
| Skeie et al. (2022) | Mortality, Causes of Death, and Predictors of Death among Patients On and Off Opioid Agonist Treatment: Results from a 19-Year Cohort Study | No direct comparison between buprenorphine and methadone |
| Soyka et al. (2006) | One-year mortality rates of patients receiving methadone and buprenorphine maintenance therapy - A nationally representative cohort study in 2694 patients | No measure of association calculated |
| Soyka et al. (2006) | Fatal poisoning in methadone and buprenorphine treated patients - Are there differences? | No measure of association calculated |
| Sun et al. (2022) | Evaluation of the Effectiveness of Buprenorphine-Naloxone on Opioid Overdose and Death among Insured Patients with Opioid Use Disorder in the United States | No direct comparison between buprenorphine and methadone |
| Thylstrup et al. (2020) | Incidence and predictors of drug overdoses among a cohort of >10,000 patients treated for substance use disorder | No direct comparison between buprenorphine and methadone |
| Vakkalanka et al. (2021) | Association between buprenorphine for opioid use disorder and mortality risk | No direct comparison between buprenorphine and methadone |
| Victor et al. (2021) | Buprenorphine Treatment Intake and Critical Encounters following a Nonfatal Opioid Overdose | No direct comparison between buprenorphine and methadone |
| Wakeman et al. (2020) | Comparative Effectiveness of Different Treatment Pathways for Opioid Use Disorder | No direct comparison between buprenorphine and methadone |

| | | |
|---|---|---|
| Walley et al. (2019) | The Contribution of Prescribed and Illicit Opioids to Fatal Overdoses in Massachusetts, 2013-2015 | No measure of association calculated |
| Walley et al. (2020) | Association between mortality rates and medication and residential treatment after in-patient medically managed opioid withdrawal: a cohort analysis | No direct comparison between buprenorphine and methadone |
| Wikner et al. (2014) | Opioid-related mortality and filled prescriptions for buprenorphine and methadone | No measure of association calculated |
| Wysowski (2007) | Surveillance of prescription drug-related mortality using death certificate data | No measure of association calculated |
| Zador et al. (2000) | Deaths in methadone maintenance treatment in New South Wales, Australia 1990-1995 | No measure of association calculated |

**Table S3**. Data extraction form

| Variable | Description |
|---|---|
| First_author | Last name of the first author |
| Journal | Journal where the article was published |
| Publication_year | Year in which the article was published |
| | **Study Characteristics** |
| Study_design | Study design (e.g., cohort study, or case-control study) |
| Setting | Country |
| Study_period | Study period |
| Data_source | Data source |
| Mean_fup_duration | Mean follow-up duration |
| Follow_up | Patient follow-up and censoring definition |
| Exposure_def | Exposure definition (e.g., intention-to-treat, as-treated, and time-varying) |
| Outcome_def | Outcome definition (e.g., ICD-9/10, DSM-IV, etc.) |
| Eligibility | Key inclusion and exclusion criteria |
| N_total | Total number of patients |
| N_bup | Number and % of treatment episodes with buprenorphine |
| N_met | Number and % of treatment episodes with methadone |
| | **Patient Characteristics at Baseline** |
| Mean_age | Mean age of patients |
| Male | % of male patients |
| Psychiatric | % of patients with any or specific psychiatric or mental health disorders |
| | **Effect Measures and Corresponding 95% CI** |
| Exposure_group | Exposure group |
| Reference_group | Reference group |
| Outcome | Specific outcome investigated |
| Occasion | Effect measure during maintenance treatment, after treatment discontinuation, or overall |
| N_deaths_bup | Number of buprenorphine-related deaths |
| PY_bup | Person-years for the buprenorphine group |
| N_deaths_met | Number of methadone-related deaths |
| PY_met | Person-years for the methadone group |
| Model | Statistical model employed |
| Confounders | Confounders included in the regression model |
| Confounding_ctrl | Techniques employed to control for confounding |
| Effect_measure | Type of effect measure reported (HR, OR, or IRR) |
| Estimate | Adjusted estimate of association |
| LCI95 | Lower limit of the 95% CI |
| UCI95 | Upper limit of the 95% CI |

**Table S4**. Full data extraction

| Description | Study 1 | Study 2 | Study 3 | Study 4 | Study 5 | Study 6 | Study 7 |
|---|---|---|---|---|---|---|---|
| Last name of the first author | Bech | Bell | Hickman | Jones | Kimber | Larney | Marteau |
| Journal where the article was published | BMC Health Services Research | Drug and Alcohol Dependence | Addiction | Drug and Alcohol Dependence | Lancet Psychiatry | Addiction | BMJ Open |
| Publication year | 2019 | 2009 | 2018 | 2022 | 2015 | 2023 | 2015 |
| Study design | Retrospective registry study | Retrospective data linkage study | Retrospective cohort study | Retrospective cohort study | Retrospective cohort study | Retrospective cohort study | Retrospective administrative data study |
| Country | Norway | New South Wales, Australia | UK | New South Wales, Australia | New South Wales, Australia | New South Wales, Australia | England and Wales |
| Study period | January 1, 2014-December 31, 2015 | April 1, 2006-December 31, 2006 | 1998-2014 | 2002-2017 | 2001-2010 | 2002-2017 | 2007-2012 |
| Data source | (a) Norwegian Cause of Death Registry (b) Norwegian Patient Registry | Pharmaceutical Drugs of Addiction System | (a) Clinical Practice Research Datalink (b) Office for National Statistics (mortality data) | Electronic Reporting and Recording of Controlled Drugs (formerly Pharmaceutical Drugs of Addiction System) | Pharmaceutical Drugs of Addiction System | Electronic Reporting and Recording of Controlled Drugs (formerly Pharmaceutical Drugs of Addiction System) | (a) National Health Service (b) Office for National Statistics 'Deaths Related to Drug Poisoning in England and Wales' |
| Mean follow-up duration | NI | NI | NI | NI | Median: 6.7 years per person (IQR: 3.7-8.4) | NI | NI |
| Patient follow-up and censoring definition | NI | NI | Until the start of next treatment episode, death, or after 12 months since termination of treatment, whichever came first | Until death; December 31, 2017; or 4 years from the last OAT episode ceasing | Until the start of next treatment episode, death, or end of follow-up, whichever came first | Until the start of next treatment episode, death, or December 31, 2017; | NI |
| Exposure definition | NI | NI | Time-varying with a 28-day grace period | Time-varying with a 7-day grace period | Time-varying with a 6-day grace period | Time-varying with a 7-day grace period | NI |

| | | | | | | | |
|---|---|---|---|---|---|---|---|
| Outcome definition (e.g., ICD-9/10, DSM-IV, etc.) | ICD-10 | ICD-10 | ICD-9 and ICD-10 | ICD-10 | ICD-10 | ICD-10 | ICD-10 |
| Key inclusion and exclusion criteria | Inclusion: All patients in the national OAT program who died between January 1, 2014 and December 31, 2015 during ongoing treatment or not more than 5 days after the last reported intake of OAT medication<br><br>Exclusion: Patients who died more than 5 days after last reported intake of OAT medication and those whose OAT status was unknown at the time of death | Exclusion: Individuals whose death was caused by methadone or buprenorphine from a source other than an Opioid Treatment Program, including those prescribed as analgesics | Inclusion: Patients who were prescribed buprenorphine or methadone for treating opioid use disorder or opioid dependence, patients who were 15-64 years of age<br><br>Exclusion: Patients who were prescribed buprenorphine or methadone for pain relief, Patients who received doses below the minimum expected for OST (i.e. < 20 mg/day methadone or < 4 mg/day buprenorphine), Patients younger than 15 or older than 64 | Inclusion: Diagnosis of opioid dependence, received OAT between August 1, 2002 and December 31, 2017 | Exclusion: Individuals who did not commence treatment, those in temporary programmes such as interstate patients, those in withdrawal programmes, and participants in clinical trials of buprenorphine because they were not necessarily given buprenorphine | Inclusion: All people prescribed OAT for the treatment of opioid dependence or opioid use disorder in the Australian state of New South Wales (NSW) between August 1, 2001 and September 30, 2018 | Exclusion: Prescriptions for detoxification or pain management, including sublingual formulations of buprenorphine, buprenorphine patches, methadone tablets, and methadone indicated as cough suppressants |
| Total number of patients | 200 | 16434 | 5935 | 45664 | 32033 | 37764 | NI |
| Number and % of treatment episodes with buprenorphine | NI | NI | 6050 (38.8%) | 16401 (35.9%) | 29206 (40.9%) | 55535 (52.4%)[$] | 2602374 (13.1%) |
| Number and % of treatment episodes with methadone | NI | NI | 9550 (61.2%) | 29263 (64.1%) | 42203 (59.1%) | 60688 (57.3%)[$] | 17333163 (86.9%) |

| | | | | | | | |
|---|---|---|---|---|---|---|---|
| Mean age of patients | 48.9 | At time of death:<br>(a) 39 (for those who died during treatment)<br>(b) 37 (for those who died while out of treatment) | NI | NI (However, median age given: 32) | NI | NI | NI |
| % of male patients | 74% | At time of death:<br>(a) 78.9% (for those who died during treatment)<br>(b) 73.1% (for those who died while out of treatment) | 68.81% | 67.7% | NI | 68.9% | NI |
| % of patients with any or specific psychiatric or mental health disorders | Psychiatric hospital admissions in past 5 years before death = 28%<br>Benzodiazepine prescription in past 1 year before death = 43%<br>Psychotropic medication prescription in past 1 year before death = 28%<br>Previous non-fatal overdose = 30% | NI | Existing self-harm history = 1.47%<br>Existing overdose history = 23.27%<br>Alcohol problems = 17.99%<br>Prison history = 5.64%<br>Homeless history = 2.21% | Hospitalizations in past 12 months before cohort entry:<br>- Self-harm: 11.1%<br>- Mental health: 21.7%<br>- Substance use disorder: 58.4% | NI | Non-drug mood disorder = 28.2%<br>Self-harm / suicide attempt = 19.7%<br>Psychiatric disorder = 11.4%<br>Substance use disorder = 49.2% | NI |
| Exposure group | Methadone | Methadone | Buprenorphine | Methadone | Methadone | Buprenorphine | Methadone |
| Reference group | Buprenorphine | Buprenorphine | Methadone | Buprenorphine | Buprenorphine | Methadone | Buprenorphine |
| Specific outcome investigated | Drug-induced cause of death | Overdose deaths | Drug-related poisoning mortality | Fatal opioid overdose | Drug-related overdose deaths | Fatal opioid overdose | Deaths related to drug poisoning |
| Effect measure during maintenance treatment, after treatment discontinuation, or overall | In treatment | (a) In treatment<br>(b) Diversion<br>(c) Overall (in treatment + diversion) | (a) On treatment (first 4 weeks)<br>(b) On treatment (4 weeks after initiation until end of treatment)<br>(c) Off treatment (first 4 weeks)<br>(d) Off treatment (4 weeks after | (a) In treatment (first 4 weeks)<br>(b) In treatment (4 weeks after initiation until end of treatment)<br>(c) Out of treatment (first 4 weeks) | (a) In treatment (first 4 weeks)<br>(b) In treatment (4 weeks after initiation until end of treatment)<br>(c) Out of treatment (first 4 weeks) | Age groups:<br>(a) < 30 years<br>(b) 30-39 years<br>(c) 40-49 years<br>(d) 50+ years<br><br>Chronic disease: | Overall (in treatment + diversion) |

| | | | | | | | |
|---|---|---|---|---|---|---|---|
| | | | discontinuation until end of treatment) | (d) Out of treatment (4 weeks after discontinuation until end of treatment) | (d) Out of treatment (4 weeks after discontinuation until end of treatment) | (e) No evidence of target chronic disease<br>(f) Circulatory disease<br>(g) Kidney disease<br>(h) Liver disease<br>(i) Respiratory disease | |
| Number of buprenorphine-related deaths | 82 | (a) 0<br>(b) 2<br>(c) 2 | (a) 1<br>(b) 4<br>(c) 8<br>(d) 6 | (a) <= 5<br>(b) 62<br>(c) 57<br>(d) 308 | (a) 2<br>(b) 29<br>(c) 18<br>(d) 125 | (a) 9<br>(b) 23<br>(c) 17<br>(d) 11<br>(e) 34<br>(f) 12<br>(g) 8<br>(h) 6<br>(i) 13 | 57 |
| Person-years for the buprenorphine group | 8487 | NI (All patients were assumed to have been in the study for 9 months [April 2006-December 2006]) | (a) 334<br>(b) 2242<br>(c) 424<br>(d) 1878 | (a) X[#]<br>(b) 50835<br>(c) 3183<br>(d) 53079 | (a) 2094.3<br>(b) 19841.9<br>(c) 1673.6<br>(d) 29565.3 | (a) 11016<br>(b) 18229<br>(c) 12901<br>(d) 6195<br>(e) 35762<br>(f) 6213<br>(g) 1794<br>(h) 1445<br>(i) 7824 | NI (Rate ratio calculated based on the number of prescriptions) |
| Number of methadone-related deaths | 109 | (a) 19<br>(b) 24<br>(c) 43 | (a) 7<br>(b) 23<br>(c) 10<br>(d) 28 | (a) 25<br>(b) 362<br>(c) 107<br>(d) 438 | (a) 18<br>(b) 151<br>(c) 10<br>(d) 206 | (a) 37<br>(b) 77<br>(c) 73<br>(d) 56<br>(e) 113<br>(f) 77<br>(g) 26<br>(h) 25<br>(i) 86 | 2366 |
| Person-years for the methadone group | 5707 | NI (All patients were assumed to have been in the study for 9 months [April 2006-December 2006]) | (a) 563<br>(b) 6924<br>(c) 620<br>(d) 3379 | (a) 3603<br>(b) 207752<br>(c) 3985<br>(d) 95166 | (a) 3343.9<br>(b) 88448.5<br>(c) 1835.5<br>(d) 43429.6 | (a) 25533<br>(b) 47208<br>(c) 32794<br>(d) 14785<br>(e) 87342<br>(f) 16123 | NI (Rate ratio calculated based on the number of prescriptions) |

| | | | | | | | |
|---|---|---|---|---|---|---|---|
| | | | | | | (g) 4654<br>(h) 3811<br>(i) 21038 | |
| Statistical model employed | Multilevel logistic regression | Pooling (to estimate rate ratio), table presented | Poisson regression | Generalized estimating equations | Poisson regression | Marginal structural models | Pooling using meta-analysis effect size calculator<br>(to estimate rate ratio), table presented |
| Confounders included in the regression model | Sex, age, Charlson Comorbidity Index score, history of non-fatal overdose and psychiatric hospital admissions in previous 5 years, and OAT duration in years | NI | Sex; age; calendar year; comorbidity score; geographical region; benzodiazepine co-prescription; gabapentoid co-prescription; number of OST patients per GP practice; number of GPs prescribing per practice; and history recorded of self-harm, overdose poisoning, alcohol problems, imprisonment or homelessness | OAT status; sex; treatment; Aboriginal and/or Torres Strait Islander status, Index of Relative Socio-Economic Disadvantage; geographical remoteness; incarceration; hospitalizations in the previous 12 months (substance use disorder, self-harm, mental health episode); mental health ambulatory outpatient activity | Sex and age | Weight-adjusted for treatment selection bias using:<br>Year, sex, geographical remoteness, Indigenous status, socio-economic disadvantage index, recency of: criminal charges, previous OAT history, most recent OAT, hospital admissions for respiratory, substance use, previous NFOD on OAT, prescriber preference<br><br>Weight adjusted for censorship using: | NI |

| | | | | | | | |
|---|---|---|---|---|---|---|---|
| | | | | | | Year, sex, geographical remoteness, indigeneity, socio-economic disadvantage index, previous OAT history, treatment and treatment × year interaction and recency of: incarceration, hospital admissions for mood and psychosis disorders, substance use and mental health ambulatory outpatient activity | |
| Techniques employed to control for confounding | Regression adjustments + Random intercepts for region included to correctly adjust the estimates for within-region correlations | NI | Propensity score technique - generated using inverse probability of treatment weights in order to balance the covariates between the two medication groups and improve model stability<br><br>Inclusion of an interaction between treatment modality and age and comorbidity | Regression adjustments | Fractional polynomial regression to find optimal functions for continuous age variable (found no significant residual confounding of the methadone-buprenorphine association due to the grouping of the age variable)<br><br>Rule-out method to establish how strong the | Marginal structural models that included inverse probability treatment and censorship weights | NI |

| | | | | | | | |
|---|---|---|---|---|---|---|---|
| | | | in the final adjusted models | | residual confounding would have to be to explain the effect sizes (concluded that the associations are unlikely to be due to unmeasured confounding) | | |
| Type of effect measure reported (HR, OR, or IRR) | Adjusted odds ratio | Crude rate ratio | Adjusted mortality incidence rate ratio (IPW adjusted + interactions) | Adjusted incidence rate ratios | Adjusted mortality rate ratio | Adjusted incidence rate ratios | Crude rate ratio |
| Estimate of association (95% CI) | 1.25 (0.63-2.48) | (a) NA<br>(b) 2.38 (0.56-10.05)<br>(c) 4.25 (1.03-17.54) | (a) 0.08 (0.01-0.48)<br>(b) 0.37 (0.17-0.79)<br>(c) 0.78 (0.36-1.66)<br>(d) 0.23 (0.12-0.48) | (a) 3.95 (0.89-17.51)<br>(b) 1.43 (0.94-2.17)<br>(c) 1.55 (0.94-2.55)<br>(d) 0.82 (0.65-1.03) | (a) 4.88 (1.73-13.69)<br>(b) 1.18 (0.89-1.56)<br>(c) 0.50 (0.29-0.86)<br>(d) 1.12 (0.96-1.31) | (a) 0.46 (0.17-1.26)<br>(b) 0.61 (0.32-1.18)<br>(c) 0.70 (0.30-1.64)<br>(d) 0.49 (0.17-1.41)<br>(e) 0.67 (0.38-1.17)<br>(f) 0.27 (0.11-0.67)<br>(g) 1.16 (0.31-4.36)<br>(h) 0.59 (0.14-2.43)<br>(i) 0.26 (0.07-0.94) | 6.23 (4.79-8.10) |

Abbreviations: NI = No Information; # = Output suppressed due to small death counts (<= 5); $ = Numbers as shown by the original study

**Table S5**. Assessment of risk of bias using ROBINS-I

| **Article** | Bech et al. (2019) | Bell et al. (2009) | Hickman et al. (2018) | Jones et al. (2022) | Kimber et al. (2015) | Larney et al. (2023) | Marteau et al. (2015) |
|---|---|---|---|---|---|---|---|
| **Bias due to confounding** | | | | | | | |
| 1.1. Is there potential for confounding of the effect of intervention in this study? | PY | Y | PY | PY | Y | PY | Y |
| 1.2. Was the analysis based on splitting participants' follow up time according to intervention received? (If N/PN, go to 1.4-1.6; If Y/PY, go to 1.3) | N | N | Y | Y | Y | Y | N |
| 1.3. Were intervention discontinuations or switches likely to be related to factors that are prognostic for the outcome? | NA | NA | PN | PN | PN | PN | NA |
| *Questions relating to BL confounding only* | | | | | | | |
| 1.4. Did the authors use an appropriate analysis method that controlled for all the important confounding domains? | Y | N | PY | PY | PN | Y | N |
| 1.5. If Y/PY to 1.4: Were confounding domains that were controlled for measured validly and reliably by the variables available in this study? | PY | NA | PY | PY | PY | PY | NA |
| 1.6. Did the authors control for any post-intervention variables that could have been affected by the intervention? | N | N | N | N | N | N | N |
| *Questions relating to BL and time-varying confounding* | | | | | | | |
| 1.7. Did the authors use an appropriate analysis method that controlled for all the important confounding domains and for time- varying confounding? | N | N | PN | PN | PN | PY | N |
| 1.8. If Y/PY to 1.7: Were confounding domains that were controlled for measured validly and reliably by the variables available in this study? | NA | NA | NA | NA | NA | PY | NA |
| **Bias in selection of participants in the study** | | | | | | | |
| 2.1. Was selection of participants into the study (or into the analysis) based on participant characteristics observed after the start of intervention? | N | N | N | N | N | N | N |
| *If N/PN to 2.1, go to 2.4:* | | | | | | | |
| 2.2. If Y/PY to 2.1: Were the post-intervention variables that influenced selection likely to be associated with intervention? | NA | NA | NA | NA | NA | NA | NA |
| 2.3. If Y/PY to 2.2: Were the post-intervention variables that influenced selection likely to be influenced by the outcome or a cause of the outcome? | NA | NA | NA | NA | NA | NA | NA |
| 2.4. Do start of follow-up and start of intervention coincide for most participants? | N | N | PY | PY | PY | PY | N |
| 2.5. If Y/PY to 2.2 and 2.3, or N/PN to 2.4: Were adjustment techniques used that are likely to correct for the presence of selection biases? | N | N | NA | NA | NA | NA | N |
| **Bias in classification of interventions** | | | | | | | |

| | | | | | | | |
|---|---|---|---|---|---|---|---|
| 3.1. Were intervention groups clearly defined? | PY | PY | PY | PY | PY | PY | PY |
| 3.2. Was the information used to define intervention groups recorded at the start of the intervention? | N | N | PY | PY | PY | PY | N |
| 3.3. Could classification of intervention status have been affected by knowledge of the outcome or risk of the outcome? | N | N | N | N | PN | N | N |
| **Bias due to deviations from intended intervention** | | | | | | | |
| 4.1. Were there deviations from the intended intervention beyond what would be expected in usual practice? | PN | PN | PN | PN | PN | PN | PN |
| 4.2. If Y/PY to 4.1: Were these deviations from intended intervention unbalanced between groups and likely to have affected the outcome? | NA | NA | NA | NA | NA | NA | NA |
| 4.3. Were important co-interventions balanced across intervention groups? | NA | NA | NA | NA | NA | NA | NA |
| 4.4. Was the intervention implemented successfully for most participants? | NA | NA | NA | NA | NA | NA | NA |
| 4.5. Did study participants adhere to the assigned intervention regimen? | NA | NA | NA | NA | NA | NA | NA |
| 4.6. If N/PN to 4.3, 4.4 or 4.5: Was an appropriate analysis used to estimate the effect of starting and adhering to the intervention? | NA | NA | NA | NA | NA | NA | NA |
| **Bias due to missing data** | | | | | | | |
| 5.1. Were outcome data available for all, or nearly all, participants? | PY | PY | N | PY | PY | PY | Y |
| 5.2. Were participants excluded due to missing data on intervention status? | PY | PN | PN | PN | PN | PN | N |
| 5.3. Were participants excluded due to missing data on other variables needed for the analysis? | N | N | PN | PN | PN | N | N |
| 5.4. If PN/N to 5.1, or Y/PY to 5.2 or 5.3: Are the proportion of participants and reasons for missing data similar across interventions? | PY | NA | Y | NA | NA | NA | NA |
| 5.5. If PN/N to 5.1, or Y/PY to 5.2 or 5.3: Is there evidence that results were robust to the presence of missing data? | PY | NA | PY | NA | NA | NA | NA |
| **Bias in measurement of outcomes** | | | | | | | |
| 6.1. Could the outcome measure have been influenced by knowledge of the intervention received? | N | N | N | N | N | N | N |
| 6.2. Were outcome assessors aware of the intervention received by study participants? | PN | PN | PN | PN | PN | PN | PN |
| 6.3. Were the methods of outcome assessment comparable across intervention groups? | Y | Y | Y | Y | Y | Y | Y |
| 6.4. Were any systematic errors in measurement of the outcome related to intervention received? | N | N | N | N | N | N | N |
| **Bias in selection of the reported result** | | | | | | | |
| Is the reported effect estimate likely to be selected, on the basis of the results, from… | | | | | | | |
| 7.1. Multiple outcome measurements within the outcome domain? | N | N | N | N | N | N | N |

| | | | | | | | |
|---|---|---|---|---|---|---|---|
| 7.2. Multiple analyses of the intervention-outcome relationship? | NI | NI | NI | NI | NI | NI | NI |
| 7.3. Different Subgroups? | N | N | N | N | PN | N | N |

Abbreviations: Y = Yes, PY = Possibly Yes, PN = Possibly No, N = No, NA = Not Applicable, NI = No Information
